# Refuting Causal Relations in Epidemiological Time Series

**DOI:** 10.1101/2023.10.01.23296395

**Authors:** Yair Daon, Kris V Parag, Amit Huppert, Uri Obolski

**Affiliations:** Azrieli Faculty of Medicine, Bar-Ilan University, Safed, Israel; Department of Epidemiology and Preventive Medicine, School of Public Health, Faculty of Medicine, Tel-Aviv University, Tel Aviv, Israel; Department of Environmental Studies, Porter School of the Environment and Earth Sciences, Faculty of Exact Sciences, Tel Aviv University, Tel Aviv, Israel; MRC Centre for Global Infectious Disease Analysis, Imperial College London, London, UK; The Bio-statistical and Bio-mathematical Unit, The Gertner Institute for Epidemiology & Health Policy Research, Sheba Medical Center, Ramat Gan, Israel

**Author notes:** These authors contributed equally to this work (Amit Huppert, Uri Obolski).

## Abstract

Causal detection is an important problem in epidemiology. Specifically in infectious disease epidemiology, knowledge of causal relations facilitates identification of the underlying factors driving outbreak dynamics, re-emergence, and influencing immunity patterns. Moreover, knowledge of causal relations can help to direct and target interventions, aimed at mitigating outbreaks. Infectious diseases are commonly presented as time series arising from nonlinear dynamical systems. However, tools aiming to detect the direction of causality from such systems often suffer from high false-detection rates. To address this challenge, we propose BCAD (Bootstrap Comparison of Attractor Dimensions), a novel method that focuses on refuting false causal relations using a dimensionality-based criterion, with accompanying bootstrap-based uncertainty quantification. We test the performance of BCAD, demonstrating its efficacy in correctly refuting false causal relations on two datasets: a model system that consists of two strains of a pathogen driven by a common environmental factor, and a real-world pneumonia and influenza incidence time series from the United States. We compare BCAD to Convergent Cross Mapping (CCM), a prominent method of causal detection in nonlinear systems. In both datasets, BCAD correctly refutes the vast majority of spurious causal relations which CCM falsely detects as causal. The utility of BCAD is emphasized by the fact that our models and data displayed synchrony, a situation known to challenge other causal detection methods. In conclusion, we demonstrate that BCAD is a useful tool for refuting false causal relations in nonlinear dynamical systems of infectious diseases. By leveraging the theory of dynamical systems, BCAD offers a transparent and flexible approach for discerning true causal relations from false ones in epidemiology and may also find applicability beyond infectious disease epidemiology.

**Author summary:** In our study, we address the issue of detecting causal relations in infectious disease epidemiology, which plays a key role in understanding disease outbreaks and reemergence. Having a clear understanding of causal relations can help us devise effective interventions like vaccination policies and containment measures. We propose a novel method which we term BCAD to improve the accuracy of causal detection in epidemiological settings, specifically for time series data. BCAD focuses on refuting false causal relations using a dimensionality-based criterion, providing reliable and transparent uncertainty quantification via bootstrapping.

We demonstrate BCAD’s effectiveness by comparing it with a prevailing causal detection benchmark, on two datasets: one involving two strains of a pathogen in a model system, and another with real-world pneumonia and influenza incidence data from the United States. BCAD considerably improves on the benchmark’s performance, in both simulations and on real-world data.

In summary, BCAD provides a transparent and adaptable method for discerning genuine causal relations from spurious ones within systems governed by nearly deterministic laws, a scenario commonly encountered in infectious disease epidemiology. Our results indicate that BCAD holds the potential to be a valuable instrument in evaluating causal links, extending its utility to diverse domains. This research contributes to the continual endeavors aimed at improving understanding of the drivers of disease dynamics.

## 1 Introduction

Revealing causal relations, known as causal discovery, is at the heart of the scientific investigation process. This problem has been tackled by many means, ranging from heuristic approaches based on scientific reasoning, such as Koch’s postulates and their extensions [1], to algorithmic approaches based on graphical models [2]. The first principled approach to causal discovery from observational time series data was presented by Granger [3]. Granger’s investigation focused on stochastic systems and, crucially, required separability in the system, i.e. that information of the cause is removed by excluding it from the analysis. Sugihara et al. [4] then suggested a complementary approach, tailored for nonlinear dynamical systems, where such separability does not hold: Convergent Cross Mapping (CCM). Initially, CCM was applied to ecological systems, but its use quickly extended to other fields, e.g. neuroscience [5], geoscience [6], and infectious disease epidemiology [7]. The latter field has particularly grown in interest since the COVID-19 pandemic, as understanding causal relations during outbreaks may facilitate implementations of interventions such as vaccination [8–11], concentrated testing efforts [12–16] and other non-pharmaceutical interventions [17].

In the field of epidemiology, CCM has been employed to seek a “unified explanation for environmental drivers of influenza that applies globally” [7]. However, the same methodology employed to arrive at this conclusion presented an unexpected and implausible causal link, suggesting that influenza might actually influence environmental drivers [18]. In response to this, the authors of the former study suggested that the authors of the latter may have misinterpreted the results generated by CCM, mistaking synchrony for causation [19].

Furthermore, another investigation applied CCM to a model involving the evolution of two strains of a pathogen within a population, which yielded notably high false-detection rates [20]. Both studies [18, 20] criticized CCM for exhibiting an elevated false-detection rate when identifying causal relations, and suggested it may not possess the level of robustness required for applications in epidemiology.

One approach to causal detection in time series that has recently gained popularity is PCMCI (Peter-Clark Momentary Conditional Independence) [21–24]. While PCMCI demonstrates success in diverse applications, its appropriateness diminishes when applied to the specific challenges common in infectious disease epidemiology. PCMCI adopts a graphical model framework for causal detection [2, 25, 26], which does not allow it to discern the direction of causality between just two observables (e.g. two pathogens, or a pathogen and an environmental driver). This limitation stems from the fundamental principles of graphical models, and cannot be overcome by providing more data. Thus, the suitability of PCMCI wanes when confronted with the specific challenges present in infectious disease epidemiology.

To facilitate causal discovery in epidemiological settings, we present BCAD: Bootstrap Comparison of Attractor Dimensions. BCAD utilizes the notion of intrinsic dimension, similarly to previous studies [27–29]. In contrast to previous approaches, BCAD focuses on refuting causal relations, rather than asserting them [2, 25]. Furthermore, while uncertainty quantification in other dimensionality based methods [29] relies on Bayesian priors and other parametric assumptions, BCAD employs a hypothesis testing scheme based simply on the bootstrap. To demonstrate BCAD’s performance, we first test BCAD on simulations of a model with two strains [20, 30], where the direction of causality is known. Then, we demonstrate BCAD ‘s performance on real-world data of pneumonia and influenza incidence [31] and environmental drivers [32] for the 48 contiguous US states. We show that for both datasets, BCAD has high success rates in refuting false causal relations, while maintaining low rates of refuting true causal relations. We also apply CCM to both datasets and show that CCM has high false-detection rates, as suggested by previous studies [18, 20]. Applying BCAD in conjunction with CCM resolves the vast majority of these false-detection events, while not interfering with CCM’s high rates of correct identification of true causal relations [4, 19].

## 2 Materials and methods

### 2.1 Causality in dynamical systems

Consider *𝒳* a compact manifold and *ϕ* : *𝒳* → *𝒳* a diffeomorphism. We call (*𝒳, ϕ*) a dynamical system. A state of the system is *ω* ∈ *𝒳*, and thus we refer to as the system’s state-space. We call a smooth random variable *X* : *𝒳* → ℝ an observable. If the current state is *ω*, the next state would be *ϕ*(*ω*); and if the current observation is *X*_*t*_ = *X*(*ω*), then the next observation would be *X*_*t*+1_ = *X*(*ϕ*(*ω*)). A time series of an observable is thus

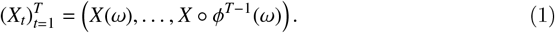

We focus our attention to systems whose states are past the initial transient phase of their dynamics. In this case *𝒳*, the state-space we consider, is an attractor [33], and every neighbourhood of every state is repeatedly visited.

Causality in dynamical systems is defined as follows [4]: Consider systems (*𝒳, ϕ*) and (*𝒴, ψ*), and a projection *π* such that *π*(*𝒴*) = and *ϕ*○*π* = *π*○*ψ* (see Supporting Information for details). We identify *𝒳* as embedded in *𝒴* and write *𝒳* ↪ *𝒴*. This setup reveals an asymmetry of separability, i.e. that information of *X* is encoded in *Y*, but not vice versa. In this case, (*𝒳, ϕ*) is known as a factor of (*𝒴, ψ*). For observables *X* : *𝒳*→ ℝ and *Y* : *𝒴*: → ℝ, we say that *X* causes *Y* and write *X* → *Y*. Thus, a factor of a system is its (non unique) cause.

For example, let us examine the relationship between absolute humidity (the pressure of water vapor in the air) and the incidence of influenza. Research has demonstrated that absolute humidity plays a role in modulating influenza transmission [34, 35]. This interaction reveals a lack of separability in the combined influenza-absolute humidity system. While the time evolution of absolute humidity can be considered irrespective of the time evolution of influenza (absolute humidity existed even before the first influenza virus emerged), the opposite does not hold: the time evolution of influenza cannot be separated from the time evolution of absolute humidity. Here, absolute humidity is a factor of a larger system that encompasses both influenza and absolute humidity, and states of the larger system contain information on both influenza and absolute humidity.

### 2.2 State-space reconstruction

In an application, we may only record time series of a single observable 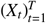. State-space reconstruction is the process of reconstructing the state space *𝒳* of a dynamical system from a finite time series of a single observable 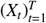. The basis to state-space reconstruction is a theorem of Takens [36] (see Supporting Information for statement). Takens’ Theorem provides a way to recover the hidden dynamics from such observed data: Generically, the state-space *𝒳* can be reconstructed from vectors of time-lagged observations *ℒ*_*X,E*_(*t*) ≔ (*X*_*t*_, …, *X*_*t*−*E*+1_), for some embedding dimension *E*. According to Takens’ Theorem, these lagged vectors form the reconstructed state-space

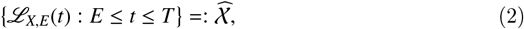

and are equivalent to states of the system. In light of Takens’ Theorem, we view the reconstructed state-space 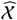 as a representative sample of the system’s state-space *𝒳*. The elements within 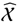 are to be regarded as authentic states of the system, comparable to states *ω* ∈ *𝒳*. Consequently, going forward, we treat 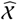 solely as a subsample extracted from *𝒳*, denoting 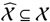.

It is important to emphasize that, as mentioned above, the conclusion of Takens’ Theorem holds only generically. Here, “generically” means that the conclusion holds on an open and dense set of *X* and *ϕ*. For example, state-space reconstruction is not possible when *X* is a constant observable; The reconstructed state-space 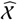 under a constant observable is a single point, even if the state-space *𝒳* is larger.

Takens’ Theorem was generalized to fractal state-spaces [37], noisy observations [38], and to deterministically [39] and stochastically forced [40] systems. We refer to such a system as a random dynamical system, as its time evolution, although governed by a deterministic law, may include a stochastic component.

According to Takens’ Theorem, it is sufficient to set the embedding dimension as *E* = 2 dim(*𝒳*) + 1. However, in practical scenarios, the value of dim(*𝒳*) is not known beforehand. Additionally, considering the curse of dimensionality, it is essential to select the smallest possible value for *E*. To address these challenges, we empirically determine the appropriate embedding dimension *E* using the False Nearest-Neighbors method [41].

Another factor that influences the reconstruction of elements in 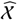 is the choice of a time lag denoted as *τ* > 0. Consequently, the elements in 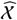 differ slightly from those described in Eq. (2). Specifically, we represent these elements as:

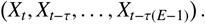

In this particular study, we opt for *τ* = 12 weeks, aligning with previous suggestions for annually forced epidemiological time series [42].

### 2.3 Estimating the Intrinsic Dimension

To estimate dim(*𝒳*) from the reconstructed state-space 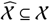, we introduce the concept of the intrinsic dimension of 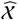, denoted as 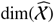. The intrinsic dimension serves as an estimation for the dimensionality of the underlying manifold or fractal *𝒳*.

The task of estimating the dimension of a manifold or fractal from a finite sample has been extensively studied, and various algorithms have been proposed and implemented for this purpose in the scikit-dimension package [43] which we employ. In our approach, we adopt the averaging of four such estimators to obtain an estimate of the intrinsic dimension. Further details on these estimators can be found in the Supporting Information.

### 2.4 Refuting causal relations with BCAD

As previously mentioned, we seek to refute false causal relations between observables of random dynamical systems. For this goal, we present BCAD: Bootstrap Comparison of Attractor Dimensions.

Consider a postulated causal relation *X* → *Y* between two observables. If this relation holds, there are two state-spaces such that *𝒳* ↪ *𝒴* and *X* : *𝒳* → ℝ, *Y* : *𝒴 →* ℝ. We therefore define a null hypothesis *H*_0_ : dim(*𝒴*) ≥dim (*𝒳*). The corresponding alternative hypothesis we pose is *H*_1_ : dim(*𝒴*) < dim(*𝒳*). To conduct hypothesis testing on *H*_0_ vs. *H*_1_, we first replace *𝒳, 𝒴*, with their corresponding reconstructed state-spaces 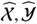. Our test statistic is 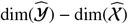, and intrinsic dimensions are calculated as described in the previous section. We quantify uncertainty in 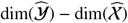 with a one-sided, bootstrap-based confidence interval, as we explain below.

To generate a single bootstrapped state space from the time series 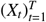, we first reconstruct the state space 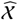, and then sample 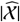 states with replacement [20, 44]. We repeat this process *B* times (we take *B* = 200 throughout this study) for both time series 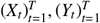. After *B* iterations, we have 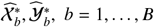 bootstrap state-spaces, sampled from 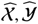, respectively. Let

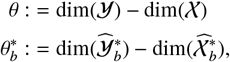

and let 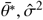 bootstrap mean and standard deviation, respectively. We construct a one-sided confidence interval for *θ* using the first-order normal approximation (the empirical bootstrap confidence interval yields nearly identical results). Since *H*_1_ is not symmetric, we find a one-sided confidence interval for *θ* at level *α* (we take *α* = 0.05 throughout this study). The rightmost edge of said confidence interval is

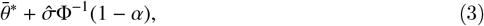

where *ϕ* is the inverse normal CDF. Thus, we reject *H*_0_ at significance level *θ* if

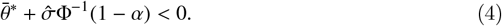

Once *H*_0_ : dim(*𝒴*) ≥ dim(*𝒳*) is rejected, BCAD has refuted *X* → *Y*.

### 2.5 Convergent cross mapping

Here we give a short overview of how CCM [4] tests *X* → *Y*, following CCM’s implementation in the package pyEDM [44], which we utilize in our experiments.

CCM is based on two components, namely cross-mapping and convergence. We start by reviewing cross-mapping, i.e. predicting one observable (e.g. *X*) from another (e.g. *Y*). We first reconstruct 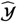 as previously described. Then, we predict observed values of *X* using Simplex Projection [45] on 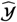 as follows: we employ a lagged vector *ℒ*_*Y,E*_(*t*^⋆^), representing unobserved data. Initially, we identify *E* + 1 lagged vectors 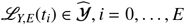, which are nearest-neighbours of *ℒ*_*Y,E*_(*t*^⋆^) in 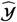 in terms of Euclidean distance. We predict 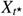 via a weighted average of 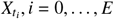, where the weight of 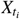 in the prediction is exponentially decreasing in the distance ‖*ℒ*_*Y,E*_(*t*^⋆^) − *ℒ*_*Y,E*_(*t*_*i*_)‖_2_. When we conduct such prediction procedure, we say that “*Y* cross-maps *X*”, or *Y* xmaps *X*.

The second component of CCM involves testing for convergence, i.e. testing if the prediction skill for *Y* xmap *X* improves with increasing amount of data. To establish convergence, we record the prediction skill of *Y* xmap *X* using all available data as *ρ*_max_. Then, we set *L*_min_ ≔ *E* + 2, as the smallest possible number of vectors required for prediction via Simplex Projection [45]. We generate *B* bootstrapped state-spaces, where we sample *L*_min_ states from 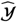 to generate each state-space. For each generated state-space, we record the prediction skill of *Y* xmap *X* using that state-space as *ρ*_min_. If *ρ*_max_ > *ρ*_min_ with statistical significance, we conclude that cross map skill has converged. We establish statistical significance when 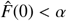 [7, 20], where 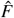 is the cumulative bootstrap distribution of *ρ*_max_ *ρ*_min_, and *α* is the predetermined significance threshold, for which we take *α* = 0.05. For visualization purposes, in our results we present the one-sided confidence interval for *ρ*_max_ − *ρ*_min_, i.e. 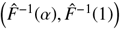.

### 2.6 Two-strain model

We use a previously published model as the data generating mechanism for our simulations [20]. In this model, two strains of a pathogen spread simultaneously in a population, and their transmission rate is modulated by an environmental driver. This two-strain model is a stochastic compartmental model that generalizes the Susceptible, Infected, Recovered (SIR) model of infectious disease epidemiology. Two main differences between the standard SIR model and the two-strain model include *β*_*i*_(*t*) the seasonally forced transmission rate and *σ*_*i j*_ the level of cross-immunity conferred to strain *i* by infection with strain *j*. The time-evolution equations of the two-strain model are the following SDEs in Itô form:

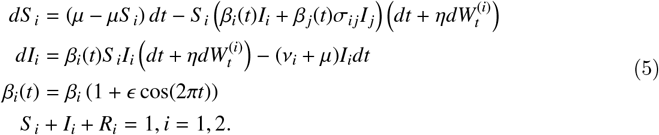

Descriptions and values of the parameters of the model are summarized in Table 1. Our choice of parameters mimics chaotic measles dynamics [30], since the parameters of the original model [20] did not display chaotic behavior (see Supporting Information for further discussion of this issue).

**Table 1.**
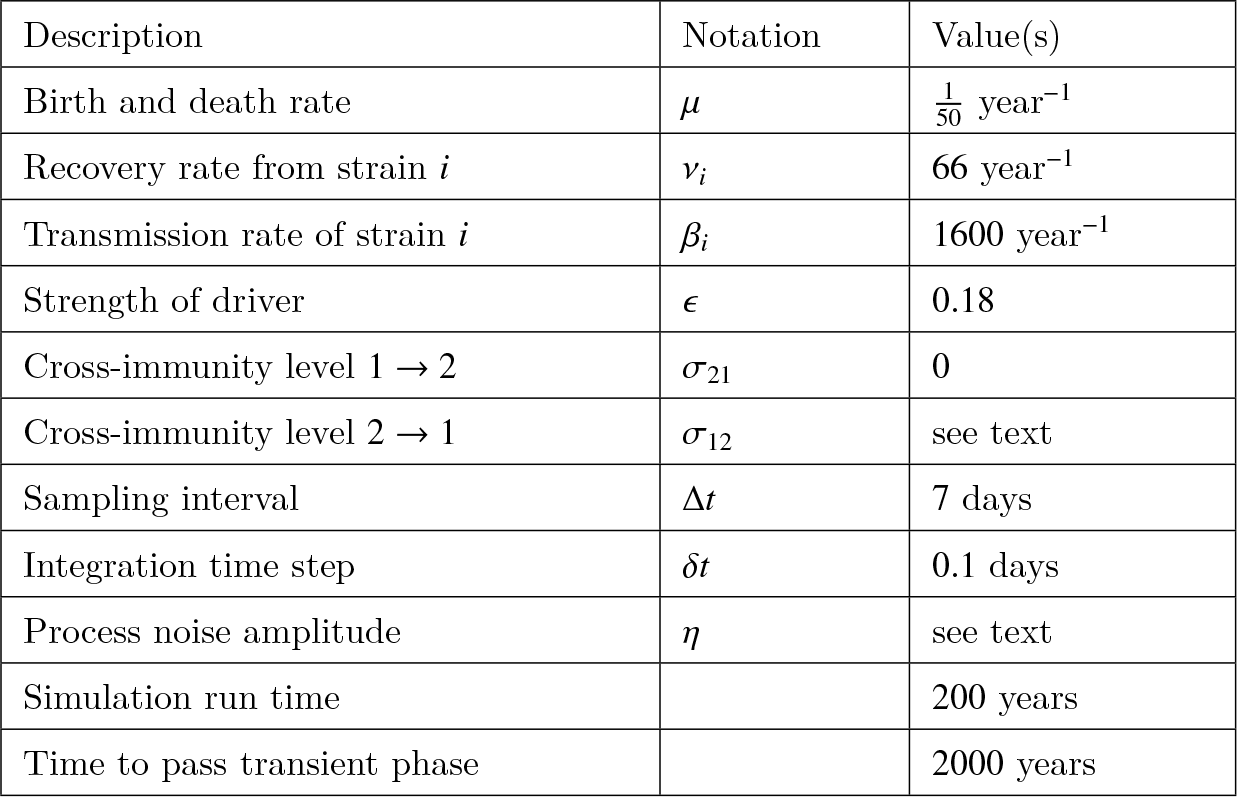
Parameters for the two-strain model.

We solve Eqs (5) with the Euler-Maruyama method with step size *δt* = 0.1 days, which is small enough to give visually satisfactory accuracy. Observation of the time series corresponding to strain *i* at some arbitrary time *t* is the cumulative incidence (incidence, henceforth) over the time interval (*t* − Δ*t, t*]:

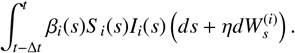

We simulate > 100 realizations for each choice of parameters with random initial conditions, and vary the process noise amplitude *η* ∈ {0, 10^−6^, …, 10^−2^} and the level of cross-immunization *σ*_12_ ∈ {0, 0.1, …, 0.9} (we noted that *σ*_12_ = 1 often results in extinction of strain 1), while keeping *σ*_21_ fixed at zero. Thus, unless *σ*_12_ = 0, strain 2 confers some degree of cross-immunity to strain 1, but not vice-versa. Consequently, the time evolution of strain 2 is a factor of the time evolution of strain 1, i.e.: 2→ 1, but 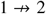.

### 2.7 Pneumonia and influenza incidence data

To examine the performance of BCAD on a real-world dataset, we used US pneumonia and influenza incidence (P&I henceforth) data, which serves as an indicator of influenza incidence [31]. Absolute humidity (the water vapor pressure in the air, AH henceforth) is known to modulate influenza transmission [34, 35], and we investigated its causal relations to P&I. We also investigated causal relations between P&I and three other environmental factors: temperature, dew-point temperature and absolute humidity (T, DP, and AH respectively). We retrieved spatio-temporal data on T and DP [32] for the relevant dates when P&I data was available, and used that to calculated AH [46, Eq. (20)].

### 2.8 Preprocessing

#### 2.8.1 Transformation of Rypdal and Sugihara

We transform epidemiological time series according to a transformation suggested by Rypdal and Sugihara [47]. This transformation is known to improve outbreak prediction when utilized within a framework of state-space reconstruction [47, 48]. For a time series of cumulative weekly incidence of an infectious disease 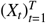, the transformation of Rypdal and Sugihara estimates the corresponding pathogen‘s reproduction number 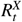 via linear regression over a running window of size *w*. Specifically:

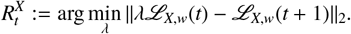

A window size of *w* = 12 weeks is a robust choice [47], and we adhere to this choice here. We transform incidence of both strains in the two-strain model, as well as P&I values.

#### 2.8.2 Filtering

Because of the detrimental effect observation noise has on intrinsic dimension calculation (the intrinsic dimension of iid noise is infinite), we filtered time series of P&I, AH, DP, and T. We did not filter simulations of the two-strain model, since those only contained process noise. We chose Singular Spectrum Analysis; a filtering method that is appropriate for filtering data arising from a random dynamical system [49, 50]. Singular Spectrum Analysis requires a threshold for removing components of the time series that correspond to observation noise, and we employ the optimal hard threshold for singular values [51] for this task. Importantly, our filtering scheme is parameter free, except for a time window required for Singular Spectrum Analysis, which we take to be 3 years — a small integer multiple of the dynamics cycle, which is a common choice [52].

## 3 Results

### 3.1 Two-strain model

First, we evaluated BCAD on synthetic data generated by the two-strain model. In Fig 2 we present “raw” incidence and corresponding estimated reproduction number for both strains and varying process noise levels *η* ∈ {0, 10^−4^, 10^−3^, 10^−2^ }. As process noise levels increase to *η* = 10^−2^, it becomes increasingly hard to distinguish dynamics from noise and hence to infer anything meaningful about the structure of causal relationships. Moreover, high levels of process noise (*η* ≥10^−2^) are qualitatively and visually different from moderate (*η* ≈10^−3^) and small (*η* ≤10^−4^) process noise levels.

**Figure 1.**
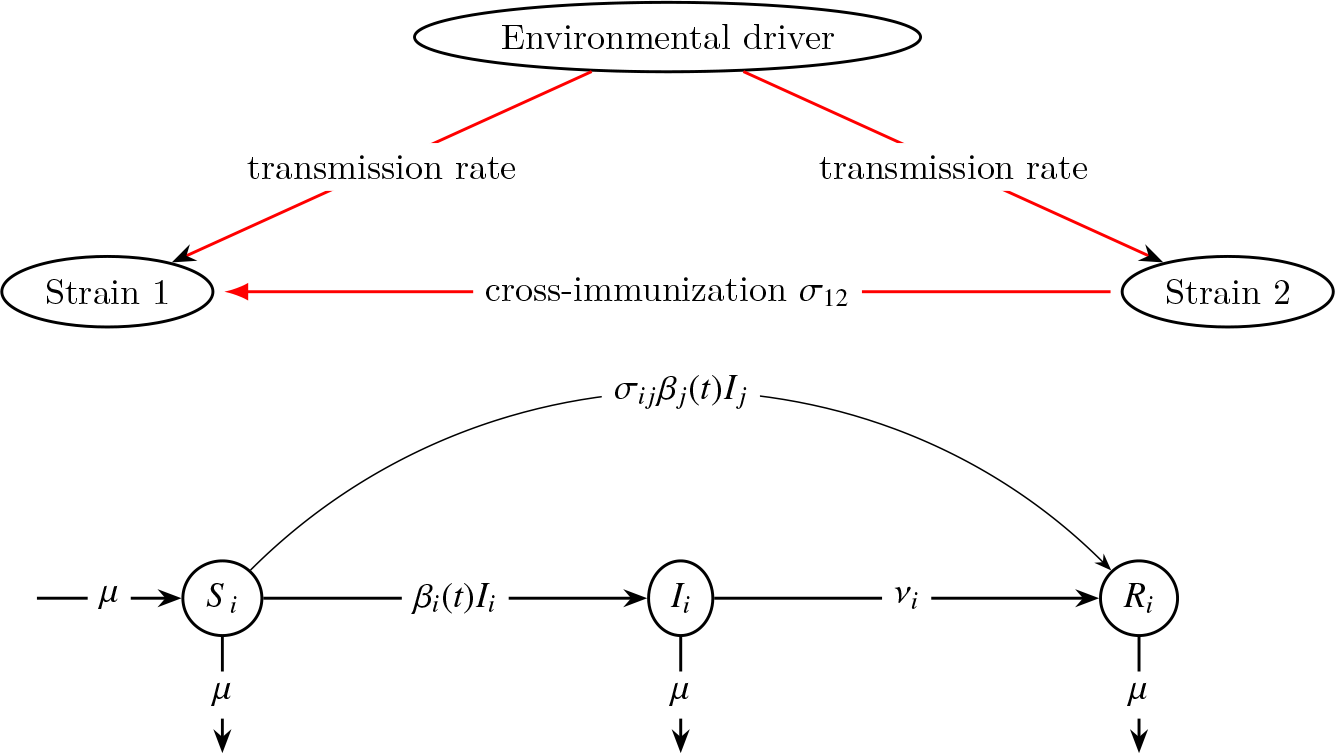
Illustration of the two-strain model. Top: a causal diagram describing how an environmental driver modulates the transmission of strains 1 and 2. Infection with strain 2 may confer cross-immunization to strain 1, but not vice-versa. Bottom: A graphical representation of the deterministic two-strain epidemic model. Rates of change are described by the quantities on top of the arrows, multiplied by the arrows’ source.

**Figure 2.**
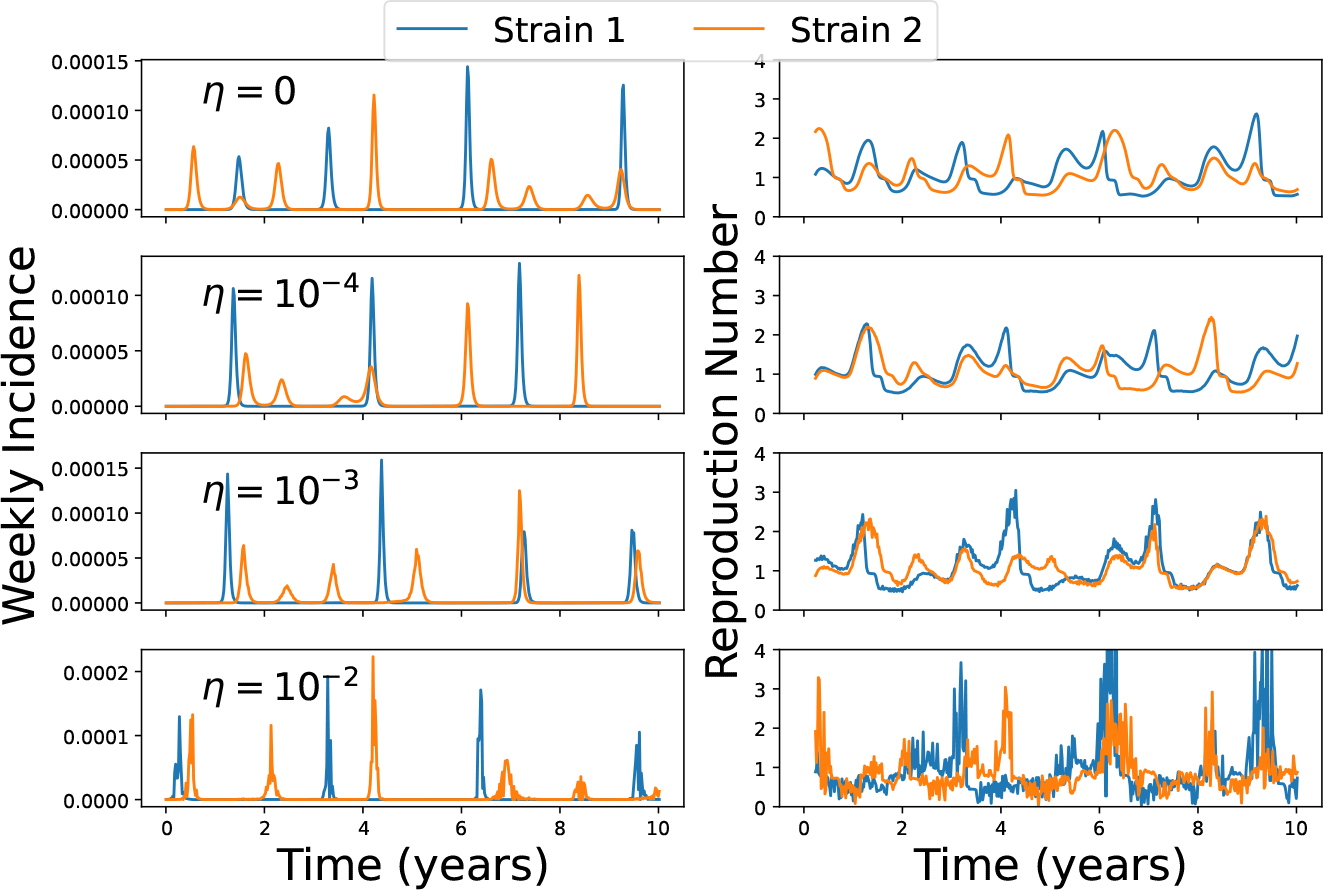
Representative time series of the two-strain model for various process noise levels. Colors represent different strains. Cross-immunization level is set to *σ*_12_ = 0.2. Each row represents a different level of process noise *η*, from no process noise (*η* = 0, top) to a high level of process noise (*η* = 0.01, bottom). Left: incidence during the sampling interval Δ*t* = 7 days. Right: Reproduction numbers for the incidence time series presented in the left panels. Note that even though the weekly incidences on the left seem constant occasionally, their underlying dynamics are revealed when we inspect the time series of their reproduction number. Furthermore, note that time series simulated with low process noise levels (*η* ≤10^−4^) are visually indistinguishable from time series simulated without process noise (*η* = 0).

As mentioned earlier, BCAD aims to refute causal relations, in contrast to CCM, which aims to discover causal relations. Since BCAD can only refute causal relations that have been posited as true by other methods, our next objective was to assess BCAD’s effectiveness when used alongside CCM—a benchmark method. Comparing the performance of BCAD and CCM is not straightforward: CCM’s null hypothesis is that no causal relation holds, i.e. 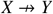. A significant increase in cross-map prediction skill implies this null hypothesis can be rejected, i.e. *X* →*Y*. In contrast, BCAD’s null hypothesis is that a causal relation holds, i.e. *X*→ *Y*. A significantly negative 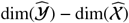 is taken as evidence to reject this hypothesis so that 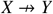 . Thus, special care has to be taken when comparing BCAD and CCM.

We simulated data, across a range of parameters with random initial conditions, from the two-strain model and tested BCAD and CCM on two causal relations: the false relation 1 →2 and the true relation 2→1 (top and bottom panels of Fig 3, respectively). We present the rejection levels of both methods, where a rejection for CCM implies that it did not identify the tested relation as causal, i.e. CCM did not reject its null hypothesis.

**Figure 3.**
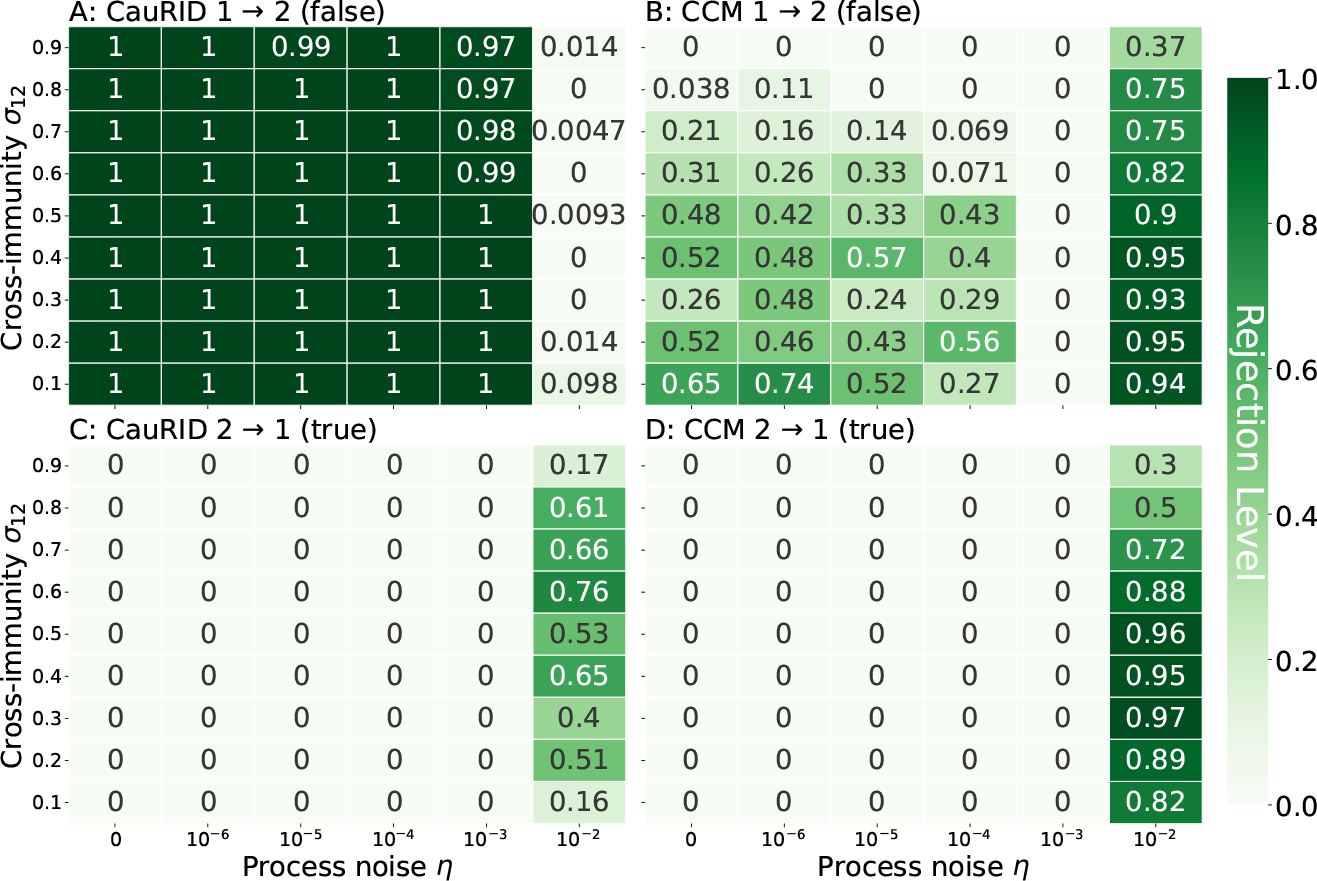
Rejection rates of BCAD (A,C) and CCM (B,D) for the two-strain model. In each panel, the X-axis represents process noise *η*, whereas the Y-axis represents the cross immunization level *σ*_12_ that strain 2 confers to strain 1 . The top panels present a relation that does not hold (1→ 2, A and B), and rejection rates should be high; the bottom panels present a relation that holds (2 →1, C and D), and rejection rates should be low. Each entry is an average of > 100 simulations. High process noise levels (*η* = 10^−2^) deteriorated the performance of both methods, so in this caption we describe results for low to moderate process noise levels *η* ≤ 10^−3^. A: BCAD correctly rejected the false relation 1 → 2 at a rate close to 100%. B: CCM rejection rates for the false relation 1 → 2 were at most 74%. Thus, CCM often did not reject the false relation 1 → 2. C, D: Both methods rejected the true relation 2 → 1 at rates close to 0%.

When cross-immunity was present (0 < *σ*_12_ < 1) and process noise was at most moderate (*η* ≤10^−3^), BCAD rejected the false relation 1 →2 at rates very close to 100% (Fig 3A), whereas CCM’s rejection rates varied substantially, ranging from 0−74% (Fig 3B). When examining the true causal relation 2→1, both BCAD and CCM had rejection rates close to 0% (Fig 3C and Fig 3D). Sufficiently high levels of process noise (*η* = 0.01) deteriorated performance for both methods, as is reasonable to expect, given the erratic behavior of time series in the lower panels of Fig 2.

In the edge case when no interaction between the strains is present (*σ*_12_ = *σ*_21_ = 0), both methods provided mediocre results. In this scenario, BCAD’s rejection rates should ideally be 0%, because the state-spaces corresponding to both strains are identical, so neither state-space will have a dimension significantly larger than the other. However, in this scenario of zero cross-immunization, BCAD refuted the false relation 2 →1 at rates as high as 33%, see Supporting Information. In contrast to BCAD, CCM’s rejection rates for the zero cross-immunization scenario should ideally be 100%, since the systems corresponding to the two pathogens are independent, and convergence of cross-map skill is not expected. However, in this scenario, CCM rejection rates were only as high as 25%, for low to moderate process noise levels (*η*≤10^−3^), see Supporting Information.

It is important to note that when BCAD is applied to raw incidence time series, its accuracy significantly decreases; in Fig 3, BCAD has almost perfect accuracy when *σ*_12_ > 0 and *η* ≤10^−3^. Performance for raw weekly incidence time series are considerably worse, see Discussion for details.

### 3.2 US pneumonia and influenza incidence

Next, we applied BCAD to a real-world dataset of weekly pneumonia and influenza incidence (P&I) and weekly averaged absolute humidity (AH), dew-point temperature (DP) and temperature (T) time series, where causal relations are known. Below, we considered the false relation that P&I incidence causes absolute humidity P&I →AH. Complementary results for the converse relation AH →P&I, as well as results for the relations DP →P&I, P&I →DP, T→P&I and P&I→T, are provided in the Supporting Information. We focused on AH →P&I since AH is a major driver of influenza transmission [34, 35]. Accurately refuting P&I →AH is important as it indicates that the method under study does not incorrectly identify a reverse causal relation as valid. We have found that CCM was frequently susceptible to this issue (see the top right panel of Fig. 3 and the right panel of Fig. 4).

**Figure 4.**
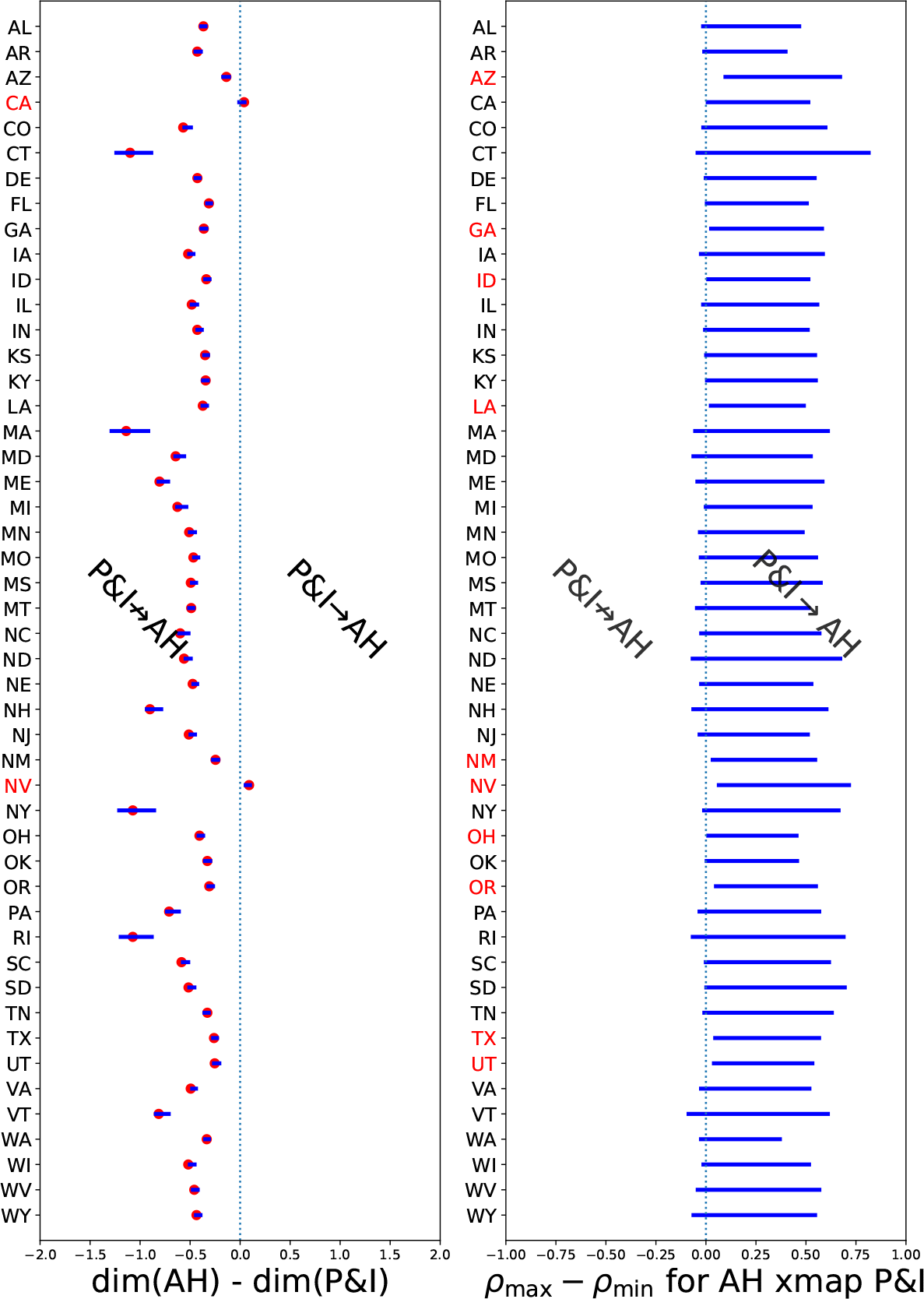
BCAD and CCM confidence intervals for the 48 contiguous states. Left: BCAD point estimates (red dots) and 90% confidence intervals (blue) for dim(AH) −dim(P&I). BCAD confidence intervals for dim(AH) −dim(P&I) were below zero for all states except for CA and NV (red). Thus, BCAD successfully refuted the false relation P&I→ AH for all but two states. We draw two-sided 90% confidence intervals for sake of presentation, even though BCAD utilizes one-sided 95% confidence intervals. Right: CCM confidence intervals for *ρ*_max−_ *ρ*_min_ when AH xmaps P&I. Confidence intervals were completely above zero for 10 states, and CCM identified the false relation P&I→ AH as true in these 10 states. Shown are 95% one-sided confidence intervals. P&I: pneumonia and influenza incidence. AH: absolute humidity.

Contrary to the previous section, where we presented rejection rates for BCAD, in this section we present BCAD confidence intervals for dim(AH)−dim(P&I). BCAD refuted P&I →AH at a 5% significance level when the 95% one-sided confidence interval of Eq (3) for dim(AH) −dim(P&I) was completely below zero. We present two-sided 90% confidence intervals for dim(AH) − dim(P&I) for aesthetic reasons and to gauge the overall precision of our estimate. However, since 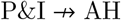, a correct inference using BCAD at a 5% significance level still corresponds to having the two-sided 90% confidence interval for dim(AH) dim(P&I) completely below zero.

BCAD correctly refuted the relation P&I→ AH for all but two states (CA and NV). Even in those two states, the CIs were close to the null (see Fig. 4). This produced a rejection rate of 46/48 = 95.8%, which is reasonable to expect with our chosen significance threshold of 5%. Furthermore, BCAD only refuted the true relation AH →P&I for NV (see Supporting Information).

Contrary to the previous section, where we presented rejection rates for CCM, in this section we present CCM confidence intervals for the cross-map skill of AH xmaps P&I. CCM asserted P&I→ AH if cross-map skill of AH xmaps P&I was significantly positive. We computed one-sided 95% confidence intervals for *ρ*_max−_ *ρ*_min_. Recall that if *ρ*_max_ −*ρ*_min_ was significantly positive, then CCM concluded that P&I−AH. Thus, a success for CCM required showing that the left edge of the corresponding confidence interval is below zero.

While CCM correctly identified the true causal relation AH→ P&I for all states (see Supporting Information), CCM incorrectly inferred P&I →AH in 10 of the 48 states (AZ, GA, ID, LA, NM, NV, OH, OR, TX and UT; see Fig 4). Thus, the false-detection rate of CCM when applied to this dataset was 10/48 ≈ 21%.

## 4 Discussion

In this study, we introduced BCAD: Bootstrap Comparison of Attractor Dimensions, a novel approach to refute causal relations by comparing intrinsic dimensions of reconstructed state-spaces. The key requirement of BCAD is for the observed time series to be governed by an underlying random dynamical system whose states have already converged to an attractor. As long as these assumptions hold (hence, Takens’ Theorem applies), BCAD can complement any causal detection scheme.

We evaluated the performance of BCAD alongside CCM — a well-known method for causal detection — to assess their respective abilities in distinguishing false and true causal relations. Our evaluation involved both simulated time series from a two-strain epidemiological model and real-world P&I time series from the US. Our results showed that BCAD correctly refuted the majority of false relations CCM mistakenly identified as true. Hence, BCAD could also be used alongside other causal detection methods, such as CCM, to avoid spurious causal discoveries.

We also noted the important role of noise filtering in causal detection for time series. Applying Singular Spectrum Analysis with a parameter-free choice of threshold was an important step in obtaining our results. Although the importance of filtering was previously mentioned [28, 29], a generic, parameter-free filtering method was not utilized. Our choice of filtering scheme reduced the possibility of cherry-picking and overfitting results by selecting from a variety of filtering parameters.

Like any method, ours is not without limitations. First, BCAD relies on time series data that is assumed to originate from observations of a random dynamical system. Additionally, BCAD assumes that the systems studied have moved past their initial transient phase; i.e. the data is expected to represent system states that are confined to the system’s attractor. When these assumptions are not met, we do not anticipate BCAD to perform effectively. Specifically, it is inadvisable to apply BCAD to scenarios where the system might have not converged to an attractor yet; e.g. incidence data related to COVID-19 during the year 2020. It is also inadvisable to apply BCAD to data of sporadic infections, since we expect the state-space of the corresponding system to be degenerate. A degenerate state-space does not allow to sensibly compare dimensionalities. Furthermore, applying BCAD to discrete data is also not advisable since state-space reconstruction is not possible for such observables, as previously explained.

Second, as with many other causal detection methods, employing BCAD may require domain knowledge. For example, when BCAD was directly applied to incidence time series from the two-strain model, its accuracy significantly decreased; long stretches of (close to) zero incidence biased dimensionality estimates. However, focusing on reproduction numbers [47, 48] extracted non-stationary features of the data (Fig. 2). Analogous issues may arise when applying BCAD to domains outside of infectious disease epidemiology, and they should be handled with relevant, domain-specific solutions.

Third, our bootstrapping scheme may have flaws that lead to erroneous inference. We conduct hypothesis testing by resampling from a reconstructed state-space, similarly to previous applications with CCM [20, 44]. Such a procedure is a hybrid between the nonparametric bootstrap (since we sample reconstructed system states with replacement) and the parametric bootstrap (since we fix a reconstructed state-space). Regardless of these considerations, we note that (a) the use of similar bootstrap schemes in CCM is widely accepted [20, 44]; (b) alternative methods for quantifying uncertainty, such as the Bayesian approach of Benkő [29] necessitate other, unverifiable underlying assumptions with their own challenges; and (c) most importantly, our method has demonstrated its efficacy on simulations and real-world data of infectious diseases time series.

By contrasting BCAD with CCM, we have demonstrated that BCAD effectively refutes causal relations falsely identified by CCM. Thus, if BCAD is used in conjunction with CCM, BCAD can increase CCM’s accuracy. Furthermore, our comparison employed epidemiological time series, akin to those for which several authors have previously reported CCM’s shortcomings [18, 20]. These time series present a significant challenge in causal detection within random dynamical systems, as the true causal relations are obscured by synchrony to a common environmental driver [19]. Synchrony is pervasive in infectious disease epidemiology, and BCAD allows us to correctly eliminate many of the false-detection events that CCM generates in the difficult scenarios where synchrony is present.

The success of BCAD in correctly refuting false causal relations in the presence of synchrony underscores its potential to significantly enhance the accuracy of causal detection. Further research may extend this improvement across various methodologies and fields of research.

## Data Availability

Data available online at https://github.com/yairdaon/BCAD

## Acknowledgments

This study was supported by the Israel Science Foundation (ISF 1286/21). Yair Daon was also supported by a post-doctoral fellowship from the Tel-Aviv University Center for Combating Pandemics.

