## Supporting Information for "Refuting Causal Relations in Epidemiological Time Series"

### 1. Definitions of attractor and factor

Definition 1 (Attractor). A compact set $\mathcal{Y}$ is an attractor if there is an open set $\mathcal{Y} \subseteq U$ such that $\phi(\bar{U}) \subseteq U$ and $\mathcal{Y} = \cap_{n \geq 0} \phi^n(U)$.

Definition 2 (Factor and extension). $(\mathcal{X}, \phi)$ is a factor of $(\mathcal{Y}, \psi)$ if there exists a surjective projection $\pi : \mathcal{Y} \to \mathcal{X}$ such that $\pi \circ \psi = \phi \circ \pi$. Conversely, $(\mathcal{Y}, \psi)$ is an extension of $(\mathcal{X}, \phi)$.

$$\begin{array}{ccc} \mathcal{Y} & \xrightarrow{\psi} & \mathcal{Y} \\ {\scriptstyle \pi}\downarrow & & \downarrow{\scriptstyle \pi} \\ \mathcal{X} & \xrightarrow{\phi} & \mathcal{X} \end{array}$$

Figure 1. $(\mathcal{X}, \phi)$ is a factor of $(\mathcal{Y}, \psi)$ via the projection map $\pi$. Conversely, $(\mathcal{Y}, \psi)$ is an extension of $(\mathcal{X}, \phi)$. We call $(\mathcal{X}, \phi)$ the cause and call $(\mathcal{Y}, \psi)$ the outcome.

The definitions above are adapted from Brin and Stuck [5]. Definition 2 is illustrated in Figure 1. As an example, consider $\mathcal{Y} = \mathcal{X} \times \mathcal{X}$ and $\psi(\omega_1, \omega_2) = (\phi_1(\omega_1), \phi_2(\omega_2))$. Then $(\mathcal{X}, \phi_1)$ is a factor of $(\mathcal{Y}, \psi)$.

### 2. Takens' Theorem

Theorem 1 ([27]). Let $\mathcal{X}$ a compact manifold, let $\phi : \mathcal{X} \to \mathcal{X}$ a $C^2$ diffeomorphism, let $X : \mathcal{X} \to \mathbb{R}$ a $C^2$ observable, and let the embedding dimension $E := 2\dim(\mathcal{X}) + 1$. Define $\mathscr{L}_{\phi,X} : \mathcal{X} \to \mathbb{R}^E$ as

$$\mathscr{L}_{\phi,X}(\omega) := \left( X(\omega), X(\phi(\omega)), \ldots, X(\phi^{E-1}(\omega)) \right). \tag{1}$$

Then generically, $\mathscr{L}_{\phi,X}$ is a $C^2$ embedding (i.e. $\mathscr{L}_{\phi,X} : \mathcal{X} \to \mathscr{L}(\mathcal{X}) \subseteq \mathbb{R}^E$ is a $C^2$ diffeomorphism). Here, "generically" means that a property holds for an open and dense set of $\phi$'s and $X$'s.

### 3. A note on our choice of parameters

The parameters initially employed by Cobey and Baskerville [8] are summarized in Table 1. We found these to be a poor choice of parameters. Below we explain why that is so. In all simulations below we took $\eta = 10^{-6}$ and $\sigma_{12} = 0.2$.

Table 1. Original parameters for the two-strain model [8]

| Description | Notation | Value(s) |
|---|---|---|
| Birth and death rate | $\mu$ | $\frac{1}{30}$ year$^{-1}$ |
| Recovery rate $i$ | $\nu$ | 5 year$^{-1}$ |
| Transmission rate of strain $i$ | $\beta_i$ | $0.25, 0.3, i = 1, 2$ year$^{-1}$ |
| Strength of environmental forcing | $\epsilon$ | 0.1 |
| Sampling interval | $\Delta t$ | 30 days |
| Simulation run time | | 1000 years |
| Year length | $\Psi$ | 360 days |
| Time to pass initial transient phase | | 2000 years |

First and foremost, we believe the choice of $\Delta t = 30$, combined with a choice of a year's length $\Psi = 360$ is not appropriate. The reason is that sampling is too synchronized with the periodicity of the time series, see Fig 2.

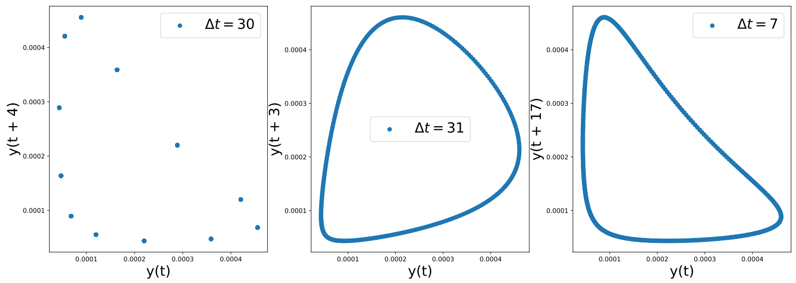


Figure 2. 2D time lag plots of time series from the two-strain model with parameters chosen according to Table 1. Time series consist of 1000 samples with sampling interval (left to right) $\Delta t = 30, 31, 7$. The time lag in the plots is $\tau \approx 120$ days (e.g. on the rightmost plot $17 \cdot 7 = 119$). The periodicity in the leftmost panel where $\Delta t = 30$ days is clearly observed.

Since $\Delta t = 30$ is not a good choice, we took $\Delta t = 7$. Still, time series generated with parameters chosen according to Table 1 are visually too synchronized. See the bottom row of Fig 3.

Lastly, time series generated from parameters of Table 1 perform too well on the S-map [26] qualitative test for deterministic chaos [26]. In Fig. 4 we see extremely high prediction skill via S-maps.

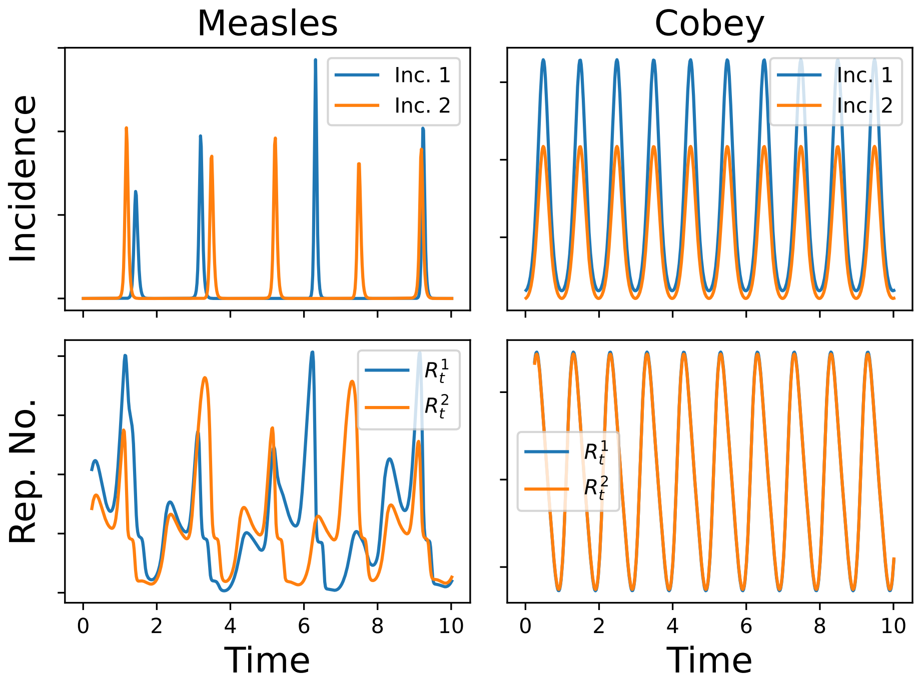


Figure 3. Left: time series of the two-strain model utilizing our choice of measles parameters. Our choice of parameters results in a seemingly chaotic behaviour. Right: Time series of the same model, with parameters chosen according to Table 1. A clear, qualitative difference can be observed between these parameter choices: almost perfect synchronization between the strains is observed.

### 4. Dimensionality Estimators

We used scikit-dimension [4] to average four dimensionality estimators: Correlation Integral [13], Maximum Likelihood Estimator [19], Method of Moments [2], and Fisher's Separability Index [1]. We did not utilize some of the estimators implemented in scikit-dimension for various reasons:

(1) Local PCA and $k$-Nearest Neighbours method [7] return only integer estimates of the intrinsic dimension, hence we have forsaken those.
(2) The DANCo [25], Two Nearest Neighbours / Minimal Neighbourhood Information [10] and Tight Localities [3] sometimes return NaN estimates. We did not use them for lack of robustness.
(3) The MiND ML algorithm of [25] always returns 1 when repeated measurements are present. Thus it does not fit for use with bootstrap.
(4) The Expected Simplex Skewness algorithm [16] takes too long to run.
(5) The Manifold-Adaptive Dimension Estimation algorithm [20] sometimes returns extremely biased results (e.g. $10^4$).

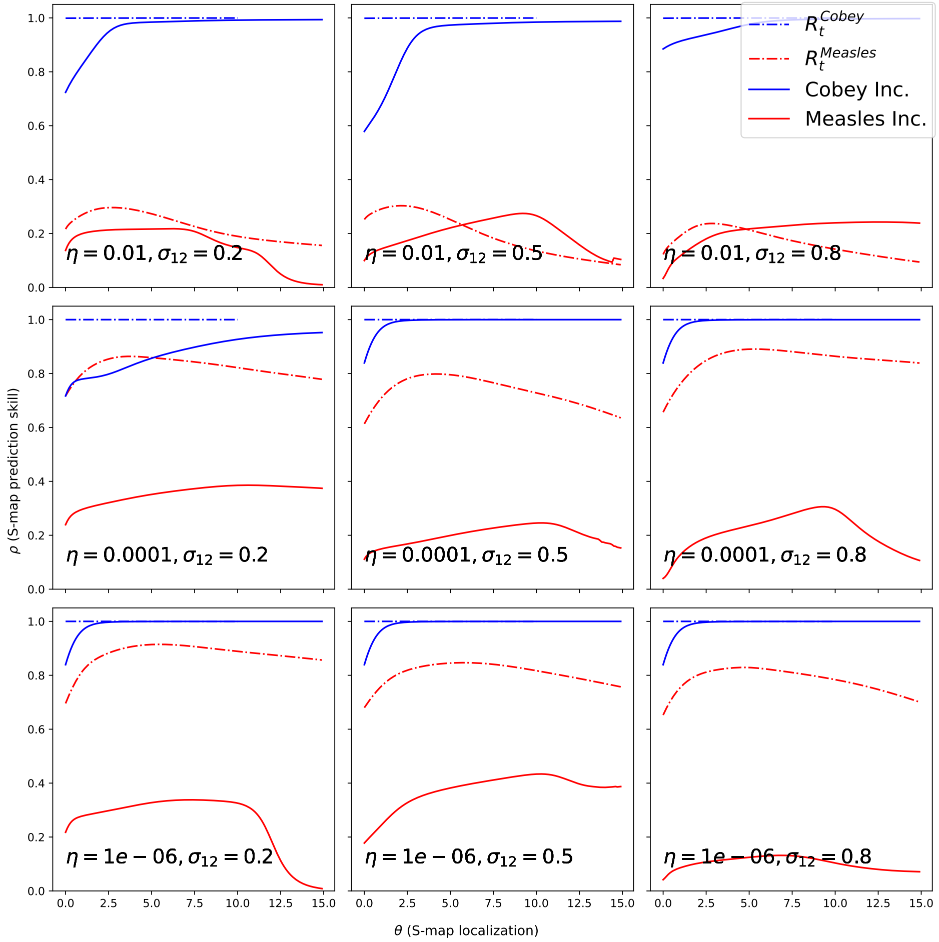


Figure 4. S-map localizations for the two-strain model. We present results for our choice of parameters, as well as the original choice of parameters [8], with and without the transformation of Rypdal and Sugihara.

### 5. Software packages

Some software packages we used: optht [9], teaspoon [22], MCSSA [30] (for our implementation of Singular Spectrum Analysis), scikit-dim [4], xarray [15], dask [24], US: The Greatest Package in the World (no citation available), shapely [12], geopandas [17], numba [18], pyEDM [23], scipy [29], numpy [14].

### 6. Repositories

Code and data. We provide an implementation of BCAD in https://github.com/yairdaon/BCAD. This repository also contains our implementation of the Singular Spectrum Analysis algorithm [6] with the optimal hard threshold [11], as well as weather [21] and P&I[28] data. Our (fast) implementation of the two-strain model with both choices of parameters can be found in https://github.com/yairdaon/fts.

Supporting Figures

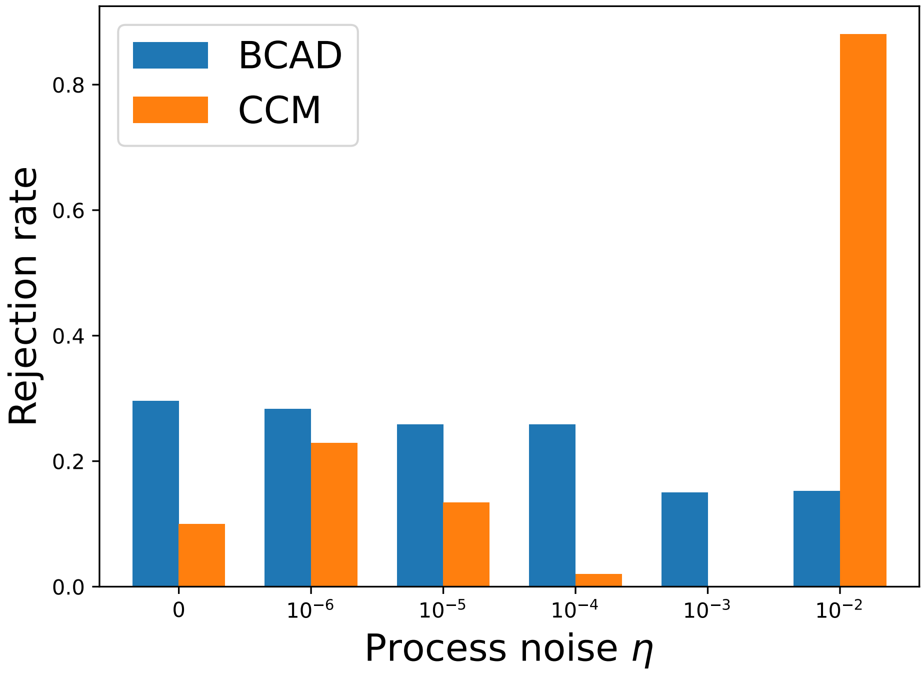


Fig. S1. Rejection rates of BCAD and CCM when $\sigma_{12} = \sigma_{21} = 0$. Rejection rates should ideally be 0% and 100% for BCAD and CCM, respectively. Note that increasing process noise level $\eta$ indeed makes BCAD approach its expected ideal behavior.

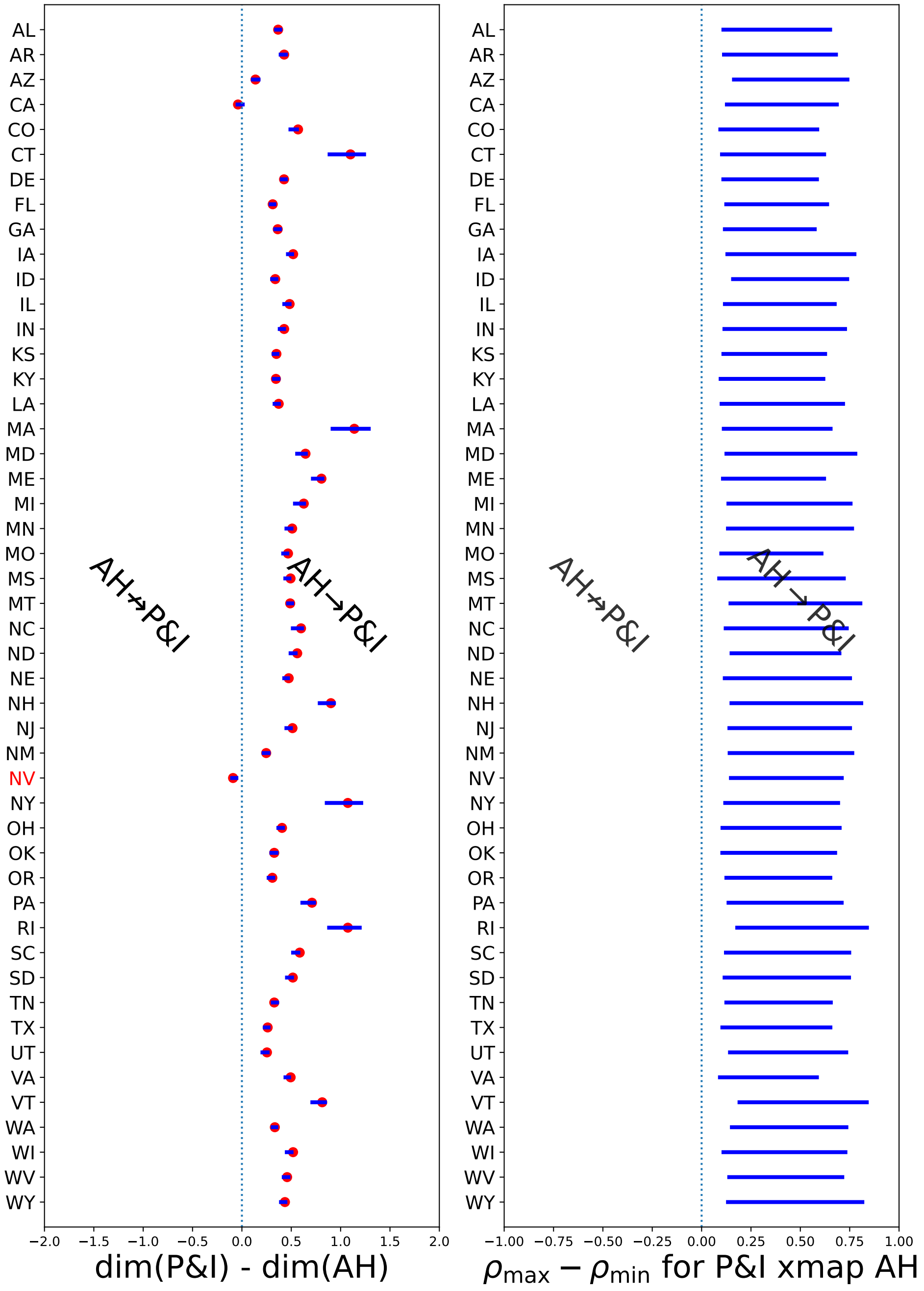


Fig. S2. Confidence intervals for BCAD and CCM testing AH → P&I.

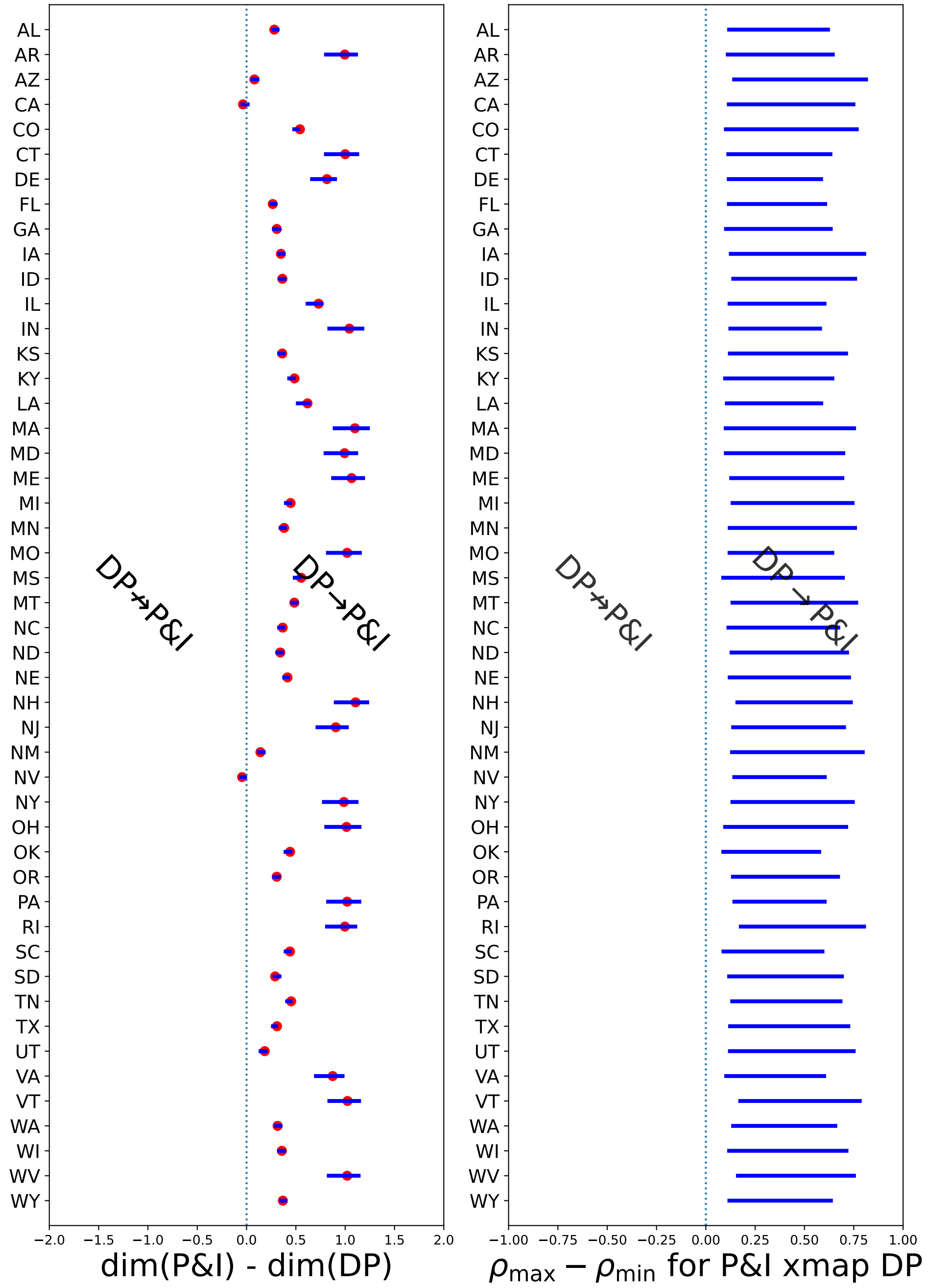


Fig. S3. Confidence intervals for BCAD and CCM testing DP → P&I.

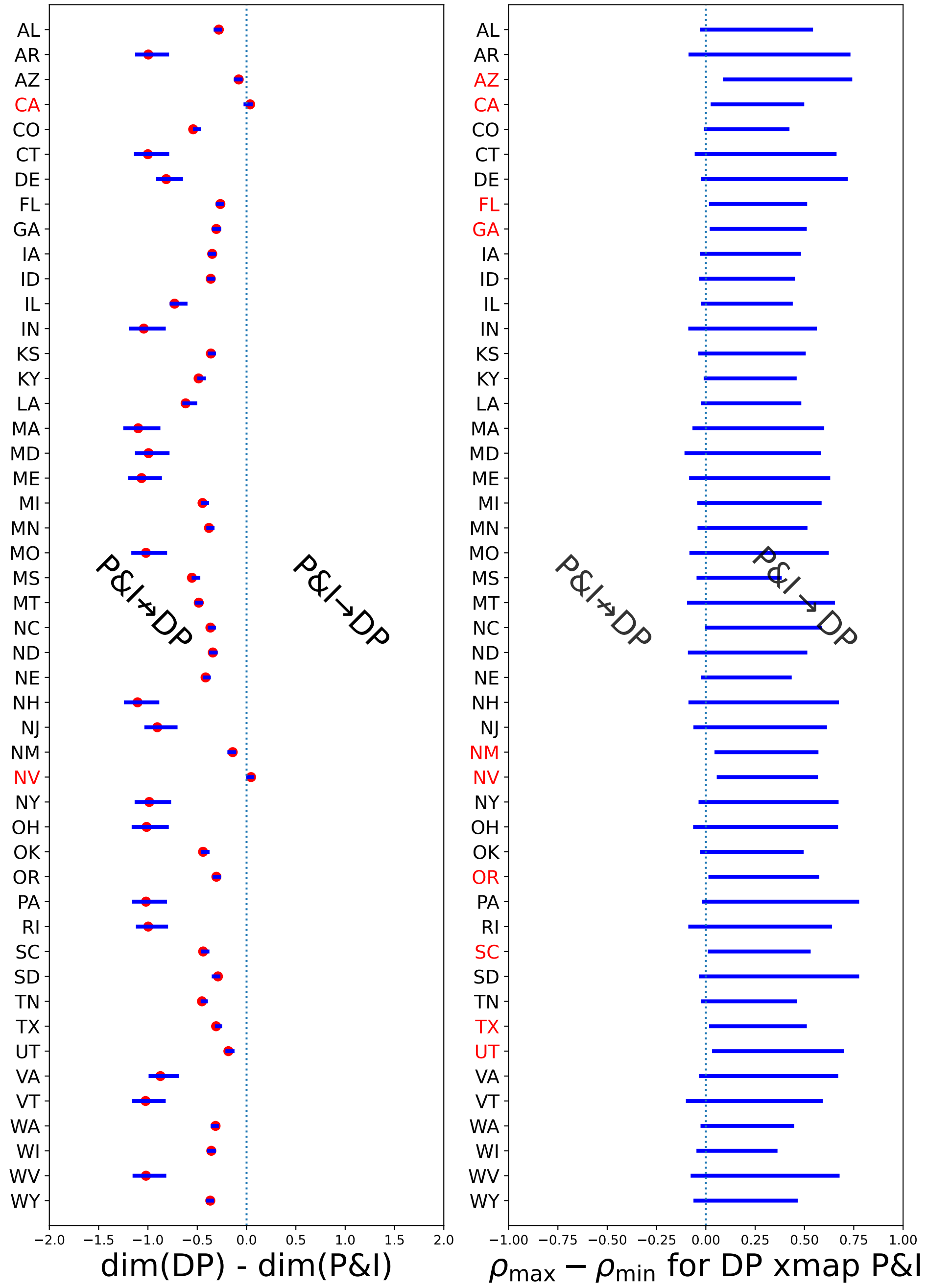


Fig. S4. Confidence intervals for BCAD and CCM testing P&I → DP.

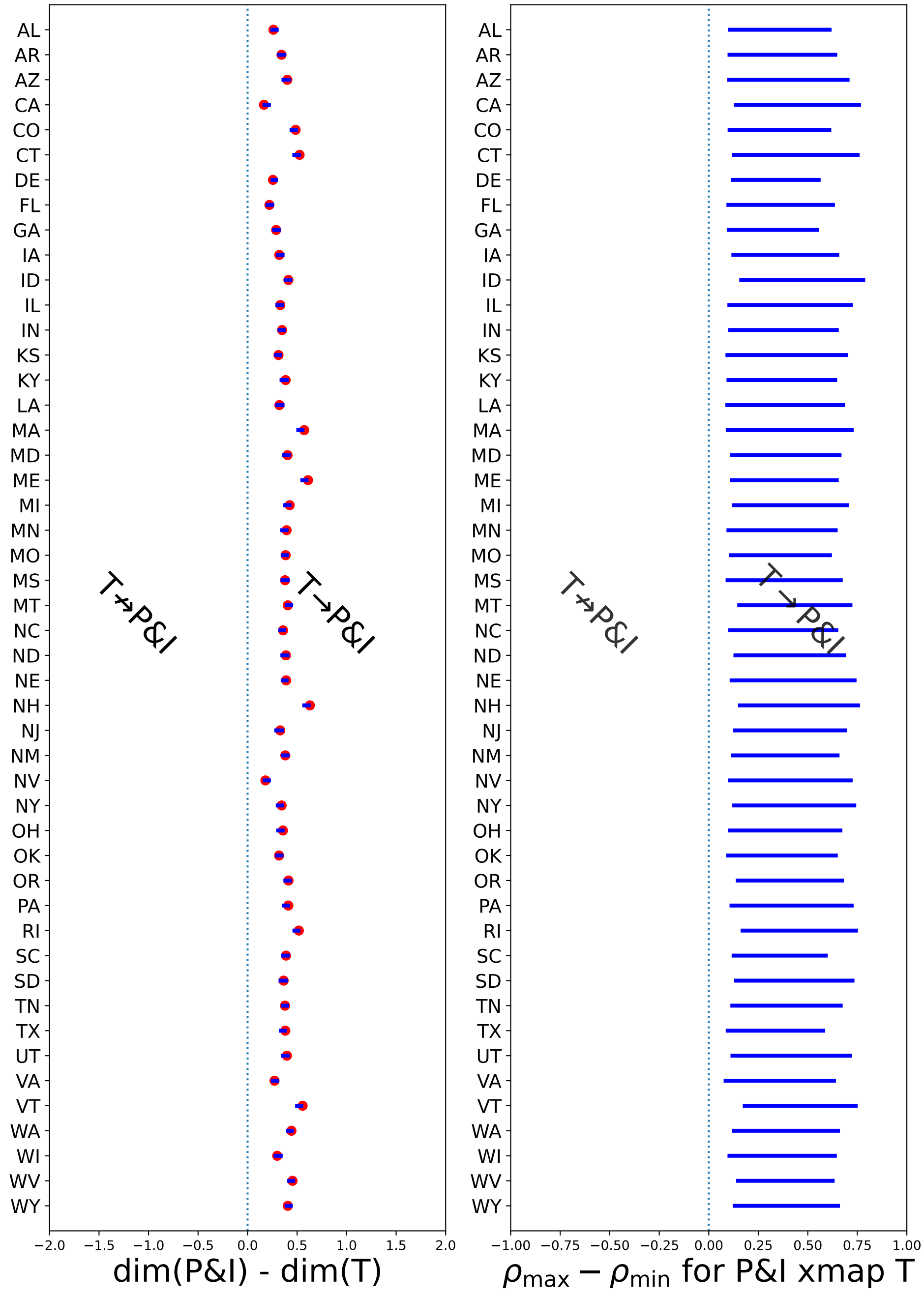


Fig. S5. Confidence intervals for BCAD and CCM testing T → P&I.

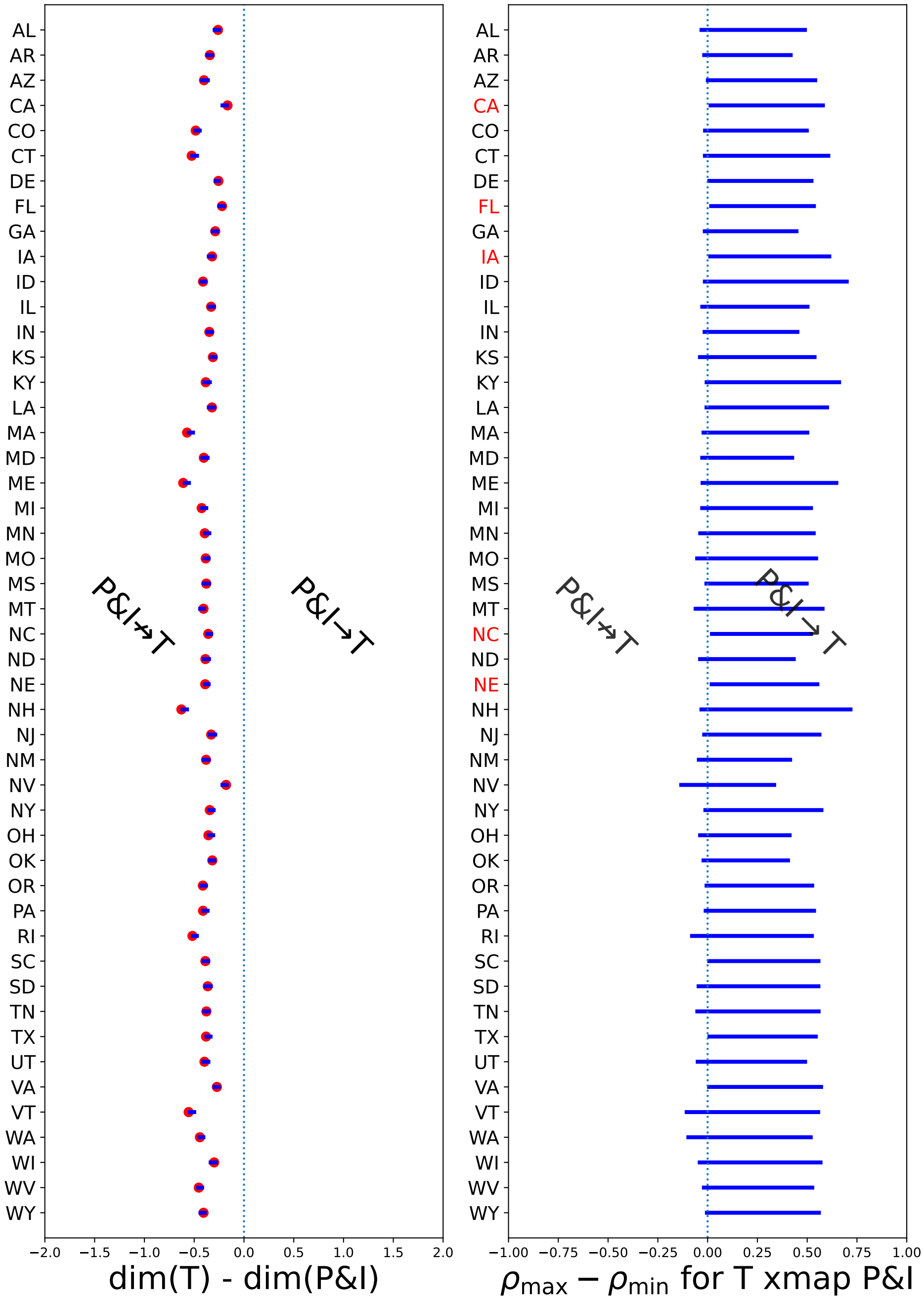


Fig. S6. Confidence intervals for BCAD and CCM testing P&I → T.

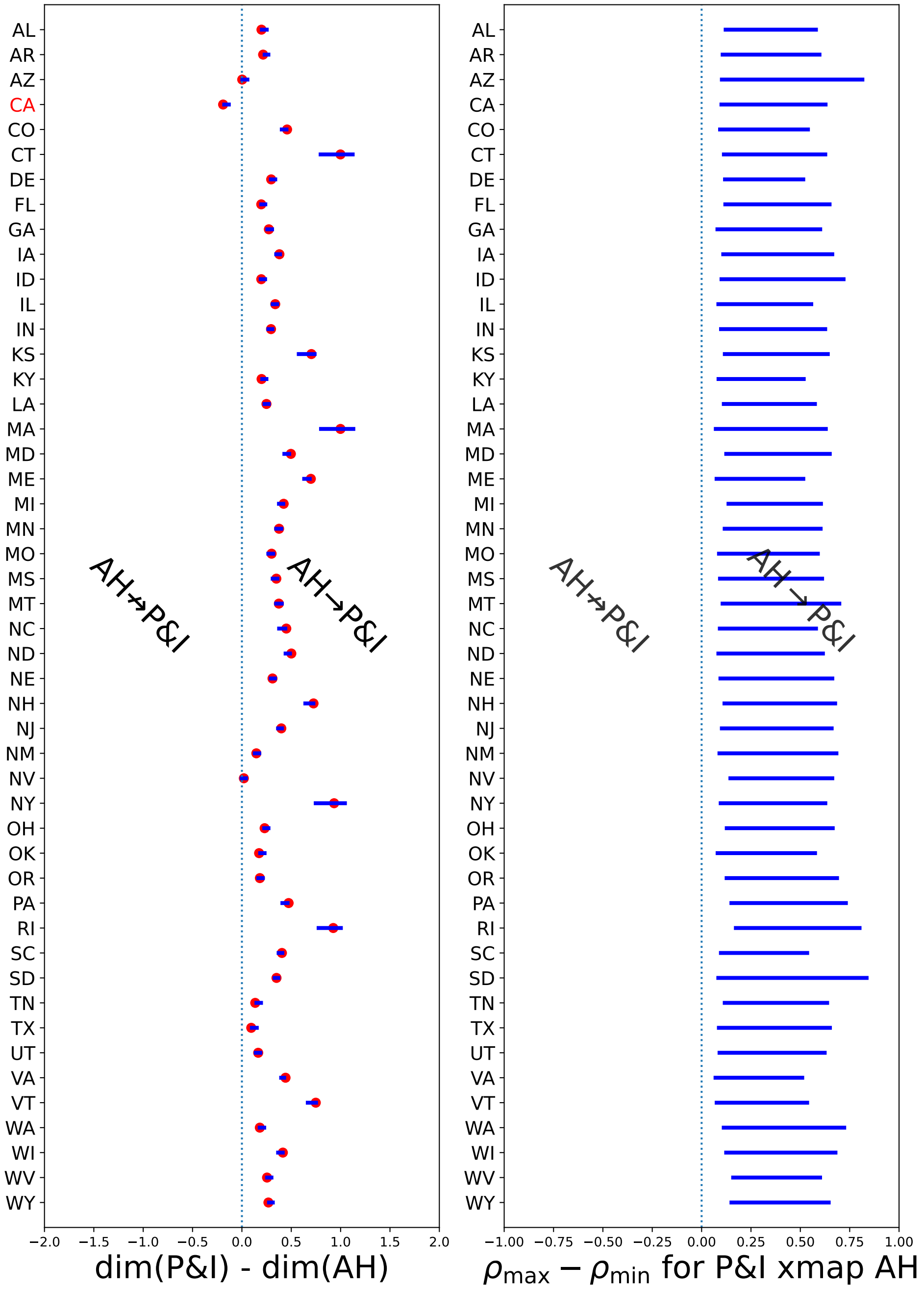


Fig. S7. Confidence intervals for BCAD and CCM testing AH → P&I, for raw P&I.

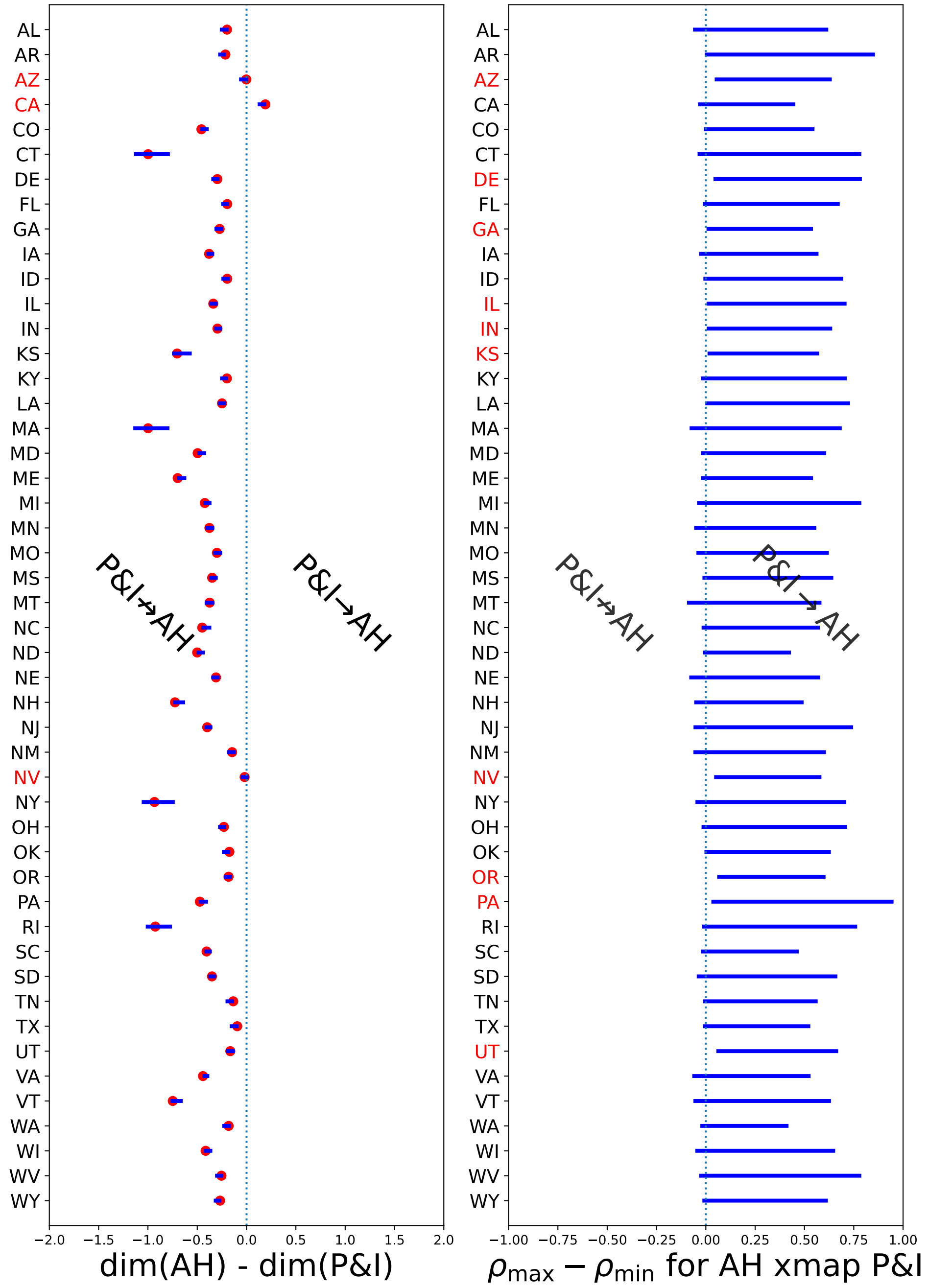


Fig. S8. Confidence intervals for BCAD and CCM testing P&I → AH, for raw P&I.

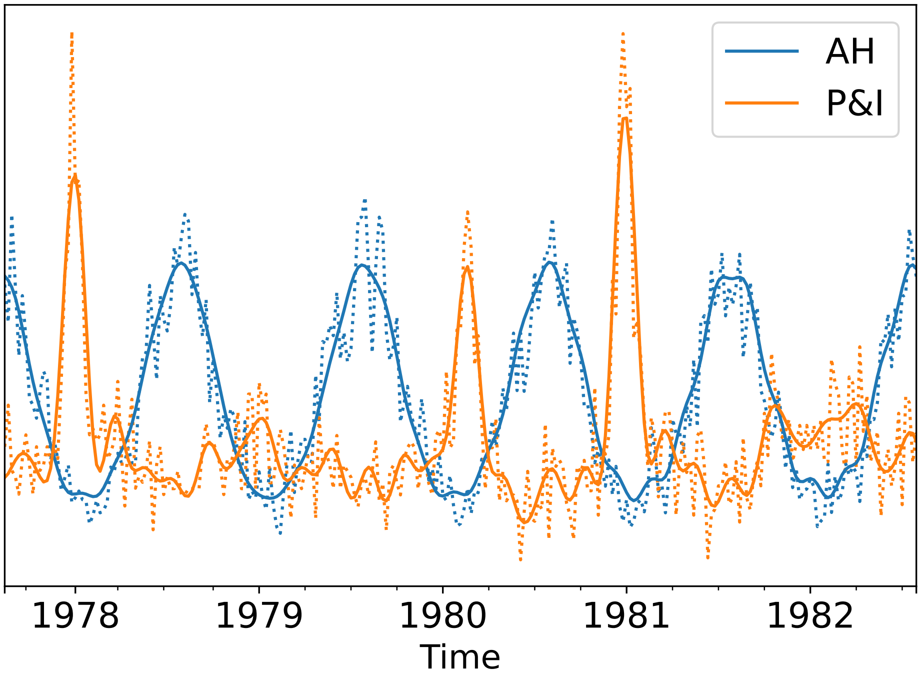


Fig. S9. Noisy and filtered time-series. Filtering via Singular Spectrum Analysis with optimal hard thresholding. Dotted lines are noisy time-series, solid lines are filtered. AH: Absolute humidity, P&I: Pneumonia and influenza incidence.

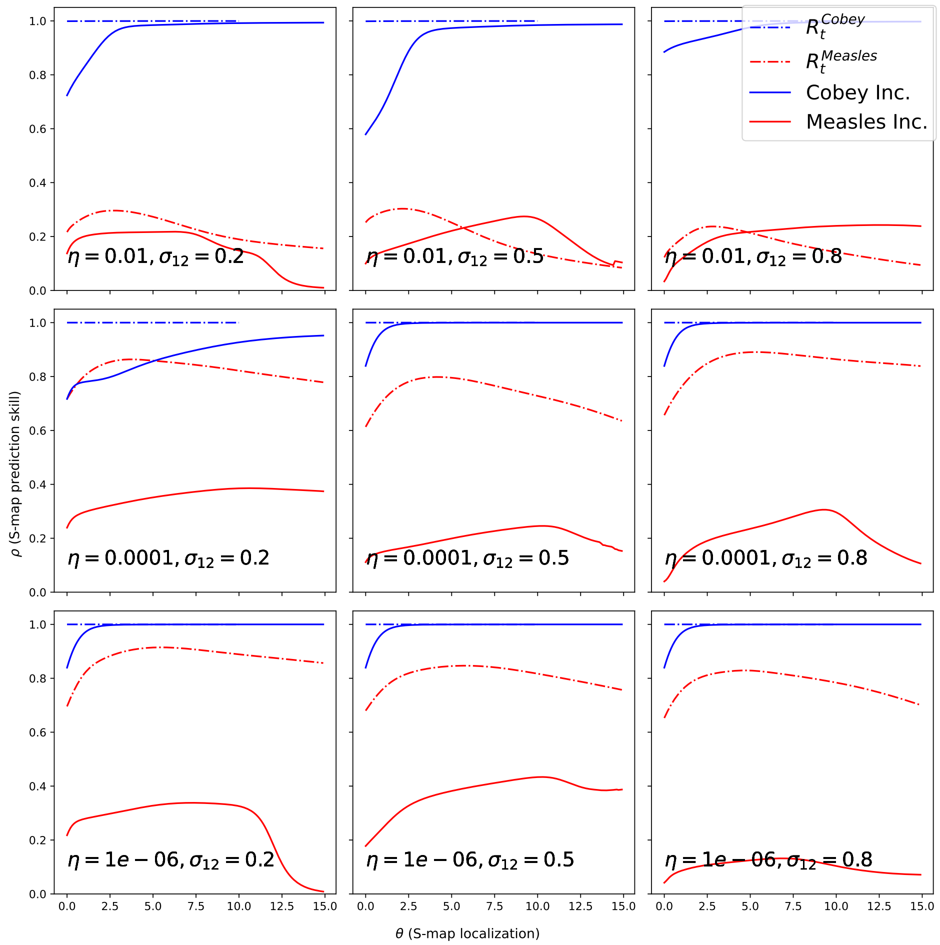


Fig. S10. Smap localization for the two-strain model. We show S-map prediction skill for the two-strain model with our choice of parameters, as well as the original choice [8]. We show localization results for ”raw” weekly incidence, as well as for its RS transformation.

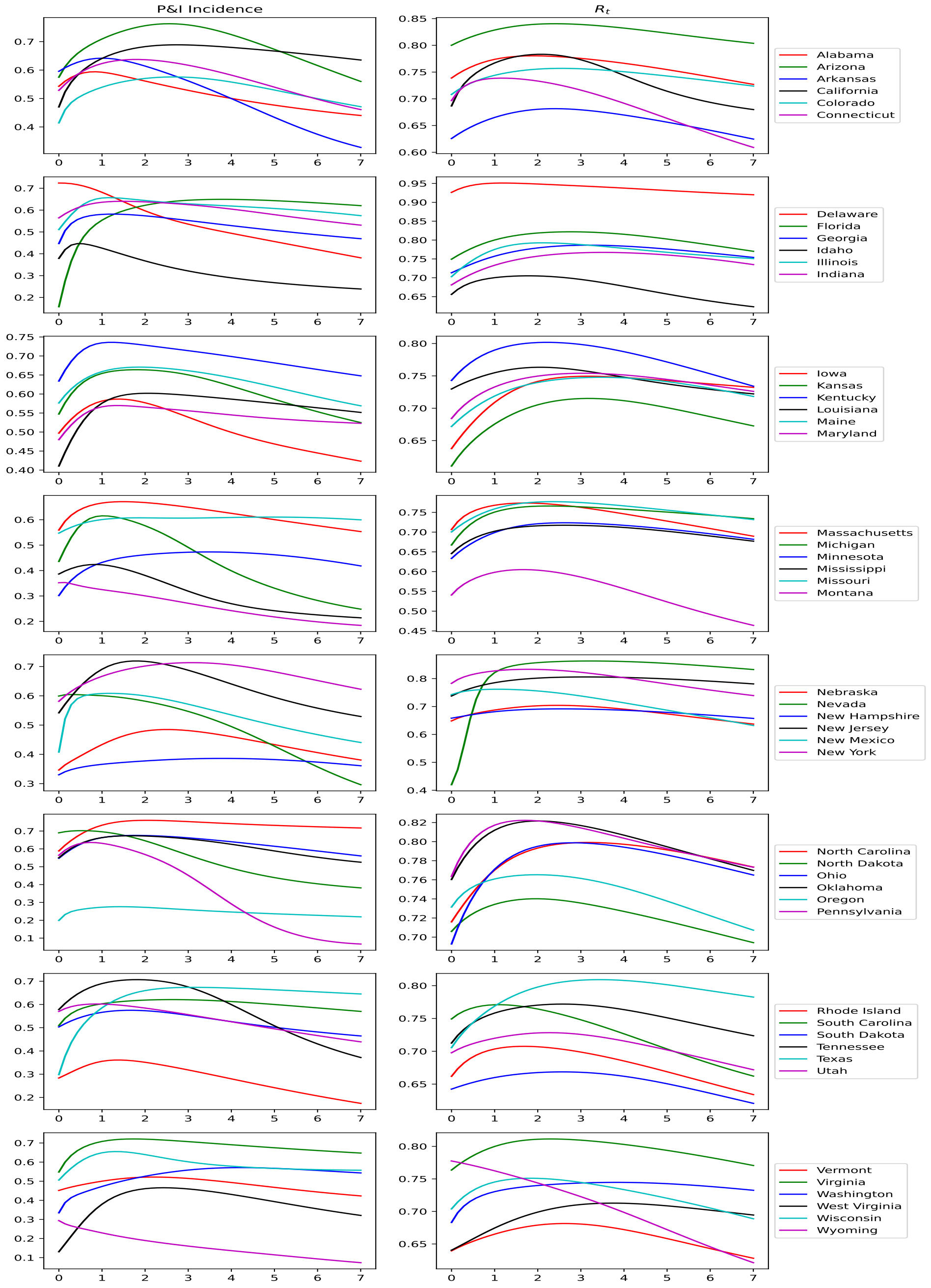


Fig. S11. Smap localization for the two-strain model. We show S-map prediction skill for 30 US states. We show S-map localization results for ”raw” weekly incidence, as well as for the reproduction numbers $R_t$.
